# Surgical window of opportunity trial reveals mechanisms of response and resistance to navtemadlin (KRT-232) in patients with recurrent glioblastoma

**DOI:** 10.1101/2024.08.12.24311893

**Authors:** Veronica Rendo, Eudocia Q. Lee, Connor Bossi, Nicholas Khuu, Michelle A. Rudek, Sangita Pal, Abigail R. N. Reynolds, Auriole C.R. Fassinou, Georges Ayoub, Emily Lapinskas, William Pisano, John Jeang, Sylwia A. Stopka, Michael S. Regan, Johan Spetz, Arati Desai, Frank Lieberman, Joy D. Fisher, Kristine Pelton, Raymond Y. Huang, Louis B. Nabors, Matthias Holdhoff, Neeraja Danda, Roy Strowd, Serena Desideri, Tobias Walbert, Xiaobu Ye, Nathalie Y. R. Agar, Stuart A. Grossman, Brian M. Alexander, Patrick Y. Wen, Keith L. Ligon, Rameen Beroukhim

## Abstract

We investigated the effectiveness of navtemadlin (KRT-232) in treating recurrent glioblastoma. A surgical window-of-opportunity trial (NCT03107780) was conducted on 21 patients to determine achievable drug concentrations within tumor tissue and examine mechanisms of response and resistance. Both 120 mg and 240 mg daily dosing achieved a pharmacodynamic impact. Sequencing of three recurrent tumors revealed an absence of *TP53*-inactivating mutations, indicating alternative mechanisms of resistance. In patient-derived GBM models, the lower range of clinically achieved navtemadlin concentrations induced partial tumor cell death as monotherapy. However, combining navtemadlin with temozolomide increased apoptotic rates while sparing normal bone marrow cells in vitro, which in return underwent reversible growth arrest. These results indicate that clinically achievable doses of navtemadlin generate significant pharmacodynamic effects and suggest that combined treatment with standard-of-care DNA damaging chemotherapy is a route to durable survival benefits.

**Statement of significance:** Tissue sampling during this clinical trial allowed us to assess mechanisms of response and resistance associated with navtemadlin treatment in recurrent GBM. We report that clinically achievable doses of navtemadlin induce pharmacodynamic effects in tumor tissue, and suggest combinations with standard-of-care chemotherapy for durable clinical benefit.

## Introduction

Over 60% of glioblastoma (GBM) express wild-type *TP53*, a tumor suppressor gene that controls cell fate decisions in response to intrinsic and extrinsic stimuli such as replicative stress and DNA damage (1). TP53 (p53) plays a critical role in preventing cancer development by activating the transcription of numerous genes involved in DNA damage repair, cell cycle arrest, and cell death by apoptosis (2). The discovery of the protein-protein interaction between TP53 and its negative regulator murine double minute homologue 2 (MDM2) has prompted the design of small molecule MDM2 inhibitors, including nutlin-3, its various chemical derivatives, and second generation agents such as KRT-232 (navtemadlin, formerly AMG 232) (3–5). As disruption of the binding of MDM2 to p53 leads to reactivation of p53 signaling in cancer cells, MDM2 inhibition has been deemed as a promising therapeutic strategy for the treatment of *TP53* wild-type tumors.

With the development of more potent and blood brain barrier-penetrant MDM2 inhibitors, new clinical trials are evaluating monotherapy and combination approaches to treat *TP53* wild-type and *MDM2*-amplified solid tumors (6–8). In the context of high-grade gliomas, MDM2 inhibition has been shown to have both *in vitro* and *in vivo* efficacy in patient-derived GBM stem cells, particularly in *TP53* wild-type tumors with *MDM2* or *MDMX* amplification (9–11). Navtemadlin is an orally bioavailable, selective small molecule inhibitor of MDM2 that blocks the protein-protein interaction between MDM2 and p53 with higher affinity than previously tested compounds (12). An initial phase I clinical trial evaluated the effect of navtemadlin in advanced solid tumors and multiple myeloma including 10 GBM patients (6), but a comprehensive assessment of brain penetration remained to be done. Therefore, we sought to perform a surgical window of opportunity trial in patients with recurrent GBM. The purpose was to ensure that at least 50% of patients attained sufficient drug concentrations (above 25 nM, the lowest concentration for which benefit in combination with radiation was seen in preclinical *in vitro* models) in contrast enhancing (CE) brain tissue. This was selected as a more useful alternative measure than a brain-to-plasma ratio cutoff based on recommendations of the Adult Brain Tumor Consortium (13). Additionally, non-contrast enhancing (NCE) brain tissue was obtained to see the difference in the concentrations attained.

## Results

### Trial outcomes

We performed a Phase 0/1 surgical window of opportunity trial (NCT03107780) of MDM2 inhibitor navtemadlin (KRT-232) in patients with GBM in association with the Adult Brain Tumor Consortium (ABTC #1604) and NCI’s Cancer Therapy Evaluation Program (CTEP) (**Fig. 1a and 1b**). In the Phase 0 (surgical window of opportunity) component, the primary objective was to determine the tumor tissue concentration and variability of navtemadlin exposure. The secondary objectives involved determining safety and toxicity as well as pharmacodynamic effects of navtemadlin on *CDKN1A*/p21 as a readout of p53 pathway engagement. The Phase 1 component, a dose escalation study of navtemadlin plus radiation in patients with newly diagnosed, O6-methylguanine DNA methyltransferase (MGMT) unmethylated, *TP53* wild-type GBM, is ongoing within Alliance for Clinical Trials in Oncology and is not reported here.

**Figure 1.**
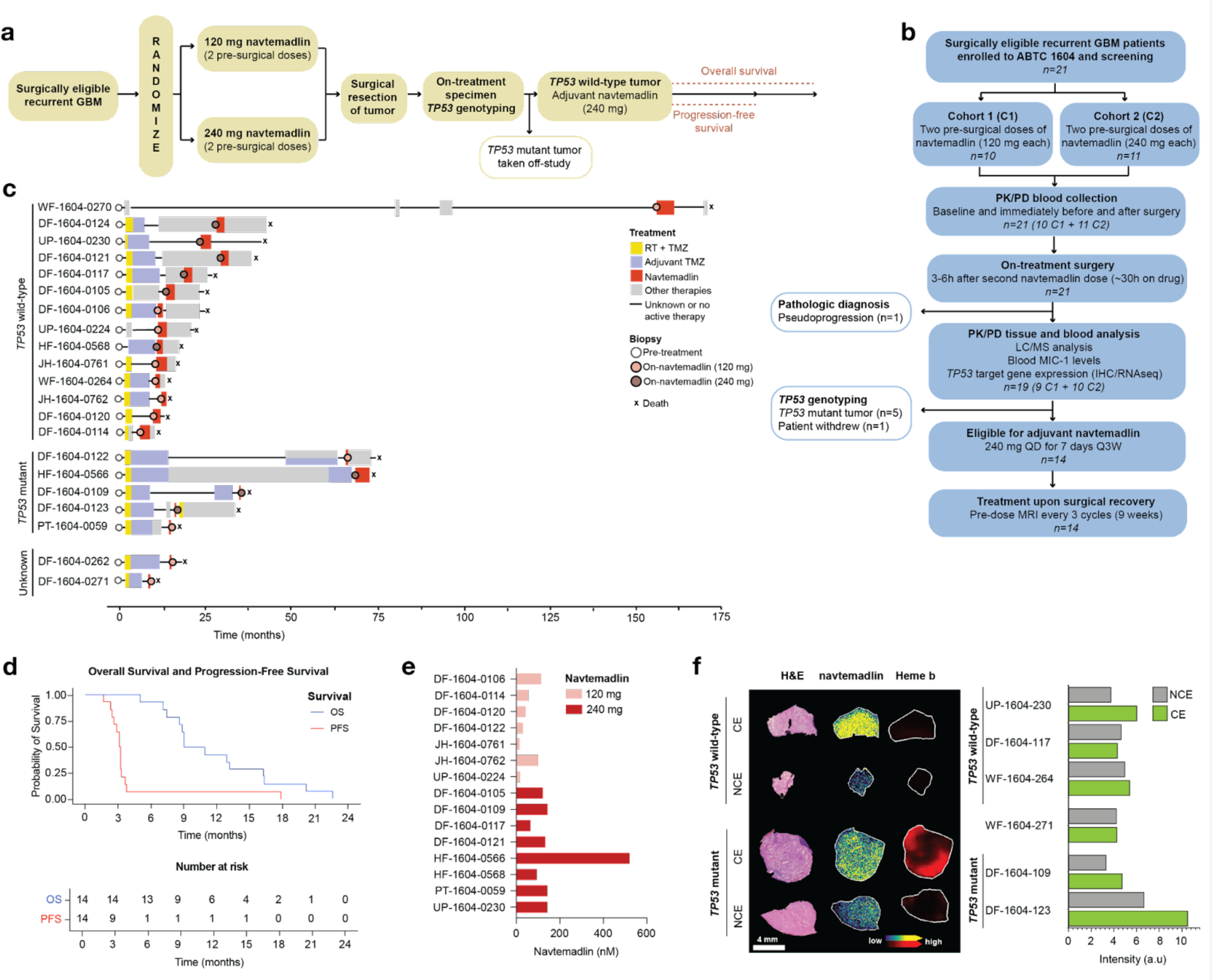
Trial schema and outcomes. **(a)** Overall schema for ABTC-1604 clinical trial and **(b)** CONSORT diagram. **(c)** Timeline for enrolled patients, indicating the duration of standard-of-care (yellow), adjuvant TMZ (purple), navtemadlin (red), as well as other (gray) or unknown (black line) treatments. The first nine patients had pre-treatment and on-navtemadlin matched biopsies available for downstream analyses. **(d)** Kaplan-Meier curves of overall survival (OS) and progression-free survival (PFS) in the patient presurgical dose cohorts. **(e)** LC-MS/MS-derived pharmacokinetic data in contrast enhancing tissue from patients receiving 120 mg (n=7) or 240 mg (n=8) navtemadlin. **(f)** Microscopy and mass spectrometry images of frozen tissue sections collected from two research subjects with *TP53* wild-type (UP-1604-0230) and *TP53* mutant (DF-1604-0123) from specimens resected in MRI contrast enhancing (CE) and non-enhancing (NCE) tumor regions. Ion images of navtemadlin (m/z 301.051) and heme b (m/z 616.178) are displayed for each specimen. Bar graph represents the average ion intensity across each tissue section, revealing higher ion intensities in specimens acquired from contrast enhancing regions compared to non-enhancing regions.

Twenty-one patients with recurrent GBM were enrolled in the surgical window of opportunity study from July 2018 to April 2020. These patients received oral navtemadlin at 120mg (n=10, minimal dose that is consistently associated with alterations in serum MIC-1, a biomarker of p53 pathway activation) or 240mg (n=11; recommended phase 2 dose as monotherapy) for two days prior surgical resection (**Fig. 1c**). Surgery was performed 3-6 h following the second administration of navtemadlin (equivalent to drug treatment for >30h). Baseline patient characteristics are noted in **Table 1**, with 18 patients diagnosed on central pathology review as GBM *IDH* wild-type and three patients as GBM *IDH* mutant as per WHO 2016 criteria. Seven patients did not receive drug after surgery (one elective withdrawal, one with no tumor/pseudoprogression on trial pathology, and five *TP53* mutant on trial pathology). The one patient with no tumor/pseudoprogression was excluded from clinical results but was retained in the safety summary. Fourteen participants continued navtemadlin following recovery from surgery at the recommended phase 2 dose of 240mg QD x 7 days Q3 weeks. Median progression-free survival (PFS) of this surgical cohort (n=14) was 3.1 months (95%CI: 2.4-3.3), and median overall survival (OS) was 10.0 months (95% CI: 7.4-16.3) (**Fig. 1d**). Overall, navtemadlin was generally well tolerated and adverse events deemed at least possibly related to navtemadlin are summarized in **Table 2**.

**Table 1.** Baseline patient characteristics.

|  | 120 mg Group (n=10) | 240 mg Group (n=10) | Total Surgical Arm (n=20) |
| --- | --- | --- | --- |
| **Age** |  |  |  |
| Median | 61 | 56 | 59 |
| Range | 40-77 | 34-71 | 34-77 |
| **Gender – No. (%)** |  |  |  |
| Male | 7 (70) | 7 (70) | 14 (70) |
| Female | 3 (30) | 3 (30) | 6 (30) |
| **Race– No. (%)** |  |  |  |
| White | 10 (100) | 10 (100) | 20 (100) |
| **KPS -- No. (%)** |  |  |  |
| 90-100 | 6 (60) | 7 (70) | 13 (65) |
| 70-80 | 2 (20) | 3 (30) | 5 (25) |
| 50-60 | 2 (20) | 0 (0) | 2 (10) |
| **Anticonvulsant – No. (%)** |  |  |  |
| Yes | 7 (70) | 7 (70) | 14 (70) |
| No | 3 (30) | 3 (30) | 6 (30) |
| **IDH -- No. (%)** |  |  |  |
| Mutant | 0 (0) | 1 (10) | 1 (5) |
| Wild type | 9 (90) | 9 (90) | 18 (90) |
| Mutant to WT | 1 (10) | 0 (0) | 1 (5) |
| **MGMT Status -- No. (%)** |  |  |  |
| Methylated | 1 (10) | 5 (50) | 6 (30) |
| Unmethylated | 9 (90) | 4 (40) | 13 (65) |
| Not Assessed | 0 (0) | 1 (10) | 1 (5) |
| **Prior No. of Relapses** |  |  |  |
| Median (range) | 1 (1-2) | 1 (1-4) | 1 (1-4) |
| **Prior Surgical Procedure – No. (%)** |  |  |  |
| Gross-total resection | 6 (60) | 4 (40) | 10 (50) |
| Sub-total resection | 4 (40) | 6 (60) | 10 (50) |
| **Prior Therapy No. (%)** |  |  |  |
| Temozolomide | 9 (90) | 9 (90) | 18 (90) |
| Other | 5 (50) | 6 (60) | 11 (55) |
| **Steroid Usage No. (%)** |  |  |  |
| Yes | 4 (40) | 2 (20) | 6 (30) |
| No | 6 (60) | 8 (80) | 14 (70) |

**Table 2.** Adverse events at least possibly related to KRT-232.

| Adverse Event<br>CTCAE v.5 | Grade 1<br>No. (%) | Grade 2<br>No. (%) | Grade 3<br>No. (%) | Grade 4<br>No. (%) | Total<br>No. (%) |
| --- | --- | --- | --- | --- | --- |
| Abdominal Pain | 1 (4.8) | 0 (0.0) | 0 (0.0) | 0 (0.0) | 1 (4.8) |
| Anemia | 5 (23.8) | 0 (0.0) | 0 (0.0) | 0 (0.0) | 5 (23.8) |
| Anorexia | 2 (9.5) | 2 (9.5) | 0 (0.0) | 0 (0.0) | 4 (19.0) |
| Atrial Fibrillation | 1 (4.8) | 0 (0.0) | 0 (0.0) | 0 (0.0) | 1 (4.8) |
| Cerebral Edema | 1 (4.8) | 0 (0.0) | 0 (0.0) | 1 (4.8) | 1 (4.8) |
| Chills | 1 (4.8) | 0 (0.0) | 0 (0.0) | 0 (0.0) | 1 (4.8) |
| Constipation | 1 (4.8) | 0 (0.0) | 0 (0.0) | 0 (0.0) | 1 (4.8) |
| Creatinine Increased | 1 (4.8) | 0 (0.0) | 0 (0.0) | 0 (0.0) | 1 (4.8) |
| Decreased Appetite | 1 (4.8) | 0 (0.0) | 0 (0.0) | 0 (0.0) | 1 (4.8) |
| Dehydration | 0 (0.0) | 1 (4.8) | 0 (0.0) | 0 (0.0) | 1 (4.8) |
| Diarrhea | 8 (38.1) | 1 (4.8) | 1 (4.8) | 0 (0.0) | 10 (47.6) |
| Dizziness | 1 (4.8) | 0 (0.0) | 0 (0.0) | 0 (0.0) | 1 (4.8) |
| Dry Mouth | 1 (4.8) | 0 (0.0) | 0 (0.0) | 0 (0.0) | 1 (4.8) |
| Dyspnea | 1 (4.8) | 0 (0.0) | 0 (0.0) | 0 (0.0) | 1 (4.8) |
| Fatigue | 2 (9.5) | 4 (19.0) | 1 (4.8) | 0 (0.0) | 7 (33.3) |
| Fecal Incontinence | 1 (4.8) | 0 (0.0) | 0 (0.0) | 0 (0.0) | 1 (4.8) |
| Flatulence | 1 (4.8) | 0 (0.0) | 0 (0.0) | 0 (0.0) | 1 (4.8) |
| Generalized Muscle Weakness | 1 (4.8) | 0 (0.0) | 0 (0.0) | 0 (0.0) | 1 (4.8) |
| Groin Rash | 0 (0.0) | 1 (4.8) | 0 (0.0) | 0 (0.0) | 1 (4.8) |
| Headache | 0 (0.0) | 2 (9.5) | 0 (0.0) | 0 (0.0) | 2 (9.5) |
| Hiccups | 0 (0.0) | 1 (4.8) | 0 (0.0) | 0 (0.0) | 1 (4.8) |
| Hyperglycemia | 1 (4.8) | 0 (0.0) | 0 (0.0) | 0 (0.0) | 1 (4.8) |
| Hyperphosphatemia | 1 (4.8) | 0 (0.0) | 0 (0.0) | 0 (0.0) | 1 (4.8) |
| Hypertension | 0 (0.0) | 1 (4.8) | 0 (0.0) | 0 (0.0) | 1 (4.8) |
| Hypoalbuminemia | 1 (4.8) | 0 (0.0) | 0 (0.0) | 0 (0.0) | 1 (4.8) |
| Hypocalcemia | 2 (9.5) | 0 (0.0) | 0 (0.0) | 0 (0.0) | 2 (9.5) |
| Hypokalemia | 4 (19.0) | 0 (0.0) | 0 (0.0) | 0 (0.0) | 4 (19.0) |
| Hyponatremia | 1 (4.8) | 0 (0.0) | 0 (0.0) | 0 (0.0) | 1 (4.8) |
| Hypophosphatemia | 1 (4.8) | 0 (0.0) | 0 (0.0) | 0 (0.0) | 1 (4.8) |
| Lymphocyte Count Decreased | 2 (9.5) | 1 (4.8) | 1 (4.8) | 0 (0.0) | 4 (19.0) |
| Nausea | 5 (23.8) | 4 (19.0) | 0 (0.0) | 0 (0.0) | 9 (42.9) |
| Neutrophil Count Decreased | 3 (14.3) | 1 (4.8) | 0 (0.0) | 1 (4.8) | 5 (23.8) |
| Pain of Skin | 0 (0.0) | 1 (4.8) | 0 (0.0) | 0 (0.0) | 1 (4.8) |
| Peripheral Sensory Neuropathy | 1 (4.8) | 0 (0.0) | 0 (0.0) | 0 (0.0) | 1 (4.8) |
| Platelet Count Decreased | 8 (38.1) | 1 (4.8) | 1 (4.8) | 1 (4.8) | 11 (52.4) |
| Stomach Pain | 1 (4.8) | 0 (0.0) | 0 (0.0) | 0 (0.0) | 1 (4.8) |
| Tinnitus | 0 (0.0) | 1 (4.8) | 0 (0.0) | 0 (0.0) | 1 (4.8) |
| Vomiting | 2 (9.5) | 2 (9.5) | 0 (0.0) | 0 (0.0) | 4 (19.0) |
| Weight Gain | 1 (4.8) | 0 (0.0) | 0 (0.0) | 0 (0.0) | 1 (4.8) |
| White Blood Cell Decreased | 8 (38.1) | 2 (9.5) | 0 (0.0) | 1 (4.8) | 11 (52.4) |

### Pharmacokinetic and pharmacodynamic analysis

The study met the pre-specified criteria of ≥25nM intra-tumor drug concentration in more than 50% of the patients for both 120mg and 240mg cohorts in CE regions of tumor. We performed liquid chromatography coupled with tandem mass spectrometry (LC-MS/MS) on CE and NCE tumor samples on 10 patients each in the 120 mg and the 240 mg cohorts. In the 120 mg cohort, 80% of patients (8/10) had drug concentrations ≥25 nM with the median being 43.1±39.6 nM in the CE tissue. In the 240 mg cohort, 100% of patients (10/10) had drug concentrations ≥25 nM with a median of 137.4±144.3 nM in the CE tissue (**Fig. 1e**).

We further evaluated the distribution of navtemadlin in both CE and NCE regions of the brain using Matrix Assisted Laser Desorption Ionization (MALDI) mass spectrometry imaging for 13 patients (**Fig. 1f; Supp. Fig. 1a**). This confirmed brain penetration, and determined higher navtemadlin concentrations in CE compared to NCE tissue for some patients. Most patients, however, exhibited similar concentrations between CE and NCE regions.

Pharmacodynamic impact was also achieved in blood samples. We assessed the fold-change in serum macrophage inhibitory cytokine (MIC-1) protein concentration by ELISA and expressed as a fold-change relative to baseline (FCB) in 10 patients in the 120 mg and the 240 mg arms. The fold variability in the two baseline (pre-dose) measurements was 0.9±0.2-fold (n=4) variability or 1.0±0.1-fold (n=6) at the 120 mg or 240 mg dose levels, respectively. Serum MIC-1 FCBs were significantly elevated approximately 24 hours after a single dose of navtemadlin. In addition there was evidence of a dose effect on serum MIC-1, as levels were 2.5-fold higher in the 240 mg cohort (9.2±4.5-FCB; n=6) than in the 120 mg cohort (3.6±2.0-FCB; n=4).

### Navtemadlin induced a pharmacodynamic response in patient tumors

To detect whether navtemadlin activates the p53 pathway in patient tumor tissue, we conducted immunohistochemistry (IHC) and bulk RNA sequencing (RNAseq) of formalin-fixed paraffin-embedded (FFPE) biopsies obtained at diagnosis (pre-treatment) and after 30 hours of drug exposure (on-treatment). Prior studies have shown MDM2 inhibitors induce *CDKN1A* (p21) RNA and protein, so we chose this p53 target as the primary tissue pharmacodynamic endpoint for the study (14). The use of paired specimens from the same patient was chosen to optimally control for potential patient-to-patient variability of baseline CDKN1A levels. On-treatment surgical tissue was obtained for all 21 patients; paired pre- and on-treatment tissue sections and evaluable RNA were available from 7 and 9 *TP53* wild-type patients, respectively (**Supp. Fig. 1b**).

Both IHC and RNAseq analyses detected p53 pathway activation in tumor tissue. In IHC analyses, we detected a significant increase in the percentage of tumor cells expressing CDKN1A with treatment (*P* = 0.01) as well as a decrease in the proliferative marker Ki67 (*P* = 0.05). Tumor p53 levels did not vary significantly with treatment (*P =* 0.11; **Fig. 2a**). We confirmed p53 pathway activation in the RNAseq data using two separate analyses. First, we compared expression levels of known transcriptional targets of p53 involved in feedback mechanisms (*MDM2*, *CCNG1* and *PPM1D*), DNA repair (*DDB2*, *RRM2B* and *PCNA*), cell cycle arrest (*BTG2*, *GADD45A* and *CDKN1A*), and apoptosis (*BAX*, *PUMA* and *TRIAP1*) (2). We detected significant upregulation of the cell cycle regulators *CDKN1A* (*P*=0.014), *BTG2* (P=0.024) and the DNA repair gene *DDB2* (*P*=0.004), as well as a trend towards upregulation of the pro-apoptosis gene *PUMA* (*P*=0.063) with navtemadlin treatment; the other targets were unchanged (**Fig. 2b; Supp. Fig. 1c**). Interestingly, *CDKN1A* expression in on-treatment biopsies also showed a positive correlation with progression-free survival (*R^2^*=0.618*; P*=0.012), suggesting that *CDKN1A* is a robust biomarker of p53 pathway activation in this patient cohort (**Fig. 2c**). These comparisons were made between samples taken from GBM patients on navtemadlin treatment and pre-treatment, meaning that most patients had been exposed to other treatments (most commonly standard of care temozolomide [TMZ] and radiation) after initial diagnosis and collection of their pre-treatment biopsies. To control for potential confounding effects of other treatments in on-treatment navtemadlin samples, we analyzed RNAseq data from an independent cohort of 25 *TP53* wild-type and *IDH* wild-type GBM tissue sample pairs obtained before and after standard-of-care treatments with temozolomide (TMZ) and radiation, profiled by the GLASS consortium [https://www.glass-consortium.org]. No upregulation of any p53 transcriptional targets was identified in these post-treatment samples (**Fig. 2b; Supp. Fig. 1d**). When comparing increases in expression values of each p53 transcriptional target between our clinical trial and GLASS control samples, we observed significant upregulation of *MDM2* (P=0.0004), *CDKN1A* (P=0.0004), *BTG2* (P=0.005)*, DDB2* (P<0.0001)*, BAX* (P=0.001) and *PUMA* (P<0.0001) in navtemadlin treated samples, suggesting a treatment specific effect. Of these canonical targets, *CDKN1A* exhibited the highest change in response to navtemadlin after ∼30 h of treatment.

**Figure 2.**
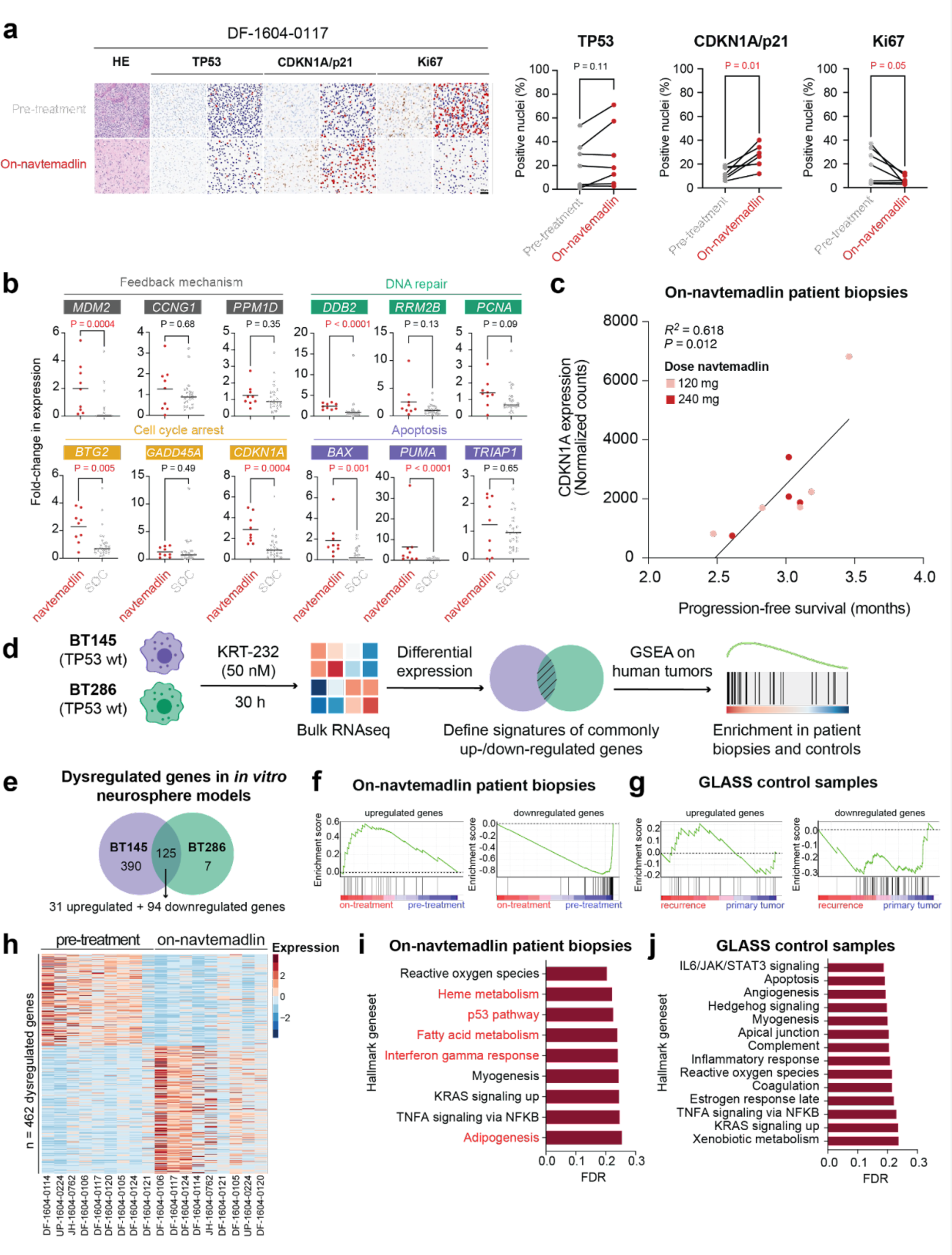
Evaluation of navtemadlin’s pharmacodynamic response in patient tumors. **(a)** Percentage of tumor cells expressing p53, CDKN1A and Ki67, as determined by immunohistochemistry and subsequent quantification of positive nuclei. Scale bar = 50 μm. **(b)** Fold-change in expression of TP53 transcriptional targets in navtemadlin patient biopsies (red; on-treatment vs pre-treatment sample) and SOC-treated GLASS control samples (gray; recurrence vs primary tumor sample). Significant gene upregulation is denoted with a red p-value. **(c)** Correlation between *CDKN1A* gene expression and progression-free survival (PFS). **(d)** *In vitro* signatures of p53 pathway activation were created by bulk RNA sequencing of two *TP53* wild-type GBM cell lines. GSEA was performed to look at enrichment in pre- and on-treatment patient biopsies and controls. **(e)** Differential expression analysis identified a total of 125 commonly dysregulated genes in BT145 and BT286 cells. **(f)** Enrichment of on-navtemadlin patient biopsies for p53 pathway signatures. **(g)** Enrichment of GLASS control samples for p53 pathway signatures. **(h)** Dysregulated genes in pre-treatment and on-treatment patient biopsies. A total of 462 genes showed differential expression with cutoffs of FDR 5% and Log_2_ Fold-change ± 1. **(i-j)** Geneset enrichment analysis for **(i)** on-navtemadlin patient biopsies and **(j)** GLASS control samples using Hallmark genesets. Pathways enriched with navtemadlin treatment and not present SOC controls are shown in red.

We detected similar pharmacodynamic effects in preclinical studies at similar drug concentrations. We performed RNA sequencing of two patient-derived *in vitro* neurosphere models (BT145 and BT286), both pre- and post-treatment with navtemadlin at an equivalent dose and time point to the clinical trial (**Fig. 2d**). We detected altered expression of 390 and 132 genes in BT145 and BT286, respectively, with 125 genes common to both (P < 0.0001) (**Fig 2e**). We used these 125 genes to construct sets of genes expected to be upregulated (n=31) and downregulated (n = 94) by treatment, and applied these gene sets to the RNAseq data from our clinical trial samples. We found on-treatment samples to be enriched for the upregulated genes (P=0.030), while pre-treatment samples were enriched for the downregulated genes (P < 0.001) (**Fig. 2f**). In contrast, analysis of GLASS samples revealed no enrichment for upregulated (P=0.727) or downregulated (P=0.647) genes in samples following standard-of-care treatment (**Fig. 2g**). We conclude that navtemadlin exhibited pharmacodynamic activity in human GBMs.

We additionally explored other pathways that were altered by navtemadlin. Using an FDR cutoff of 5%, we detected a total of 462 significantly dysregulated genes (**Fig. 2h**). Among the 50 MSigDB hallmark genesets (15), none reached an FDR threshold of 5%. The most significantly altered gene sets reflected regulation of reactive oxygen species (ROS) and cell metabolism, with the hallmark p53 pathway ranking third (FDR=22.4%; **Fig. 2i**). However, we did not observe dysregulation of the p53 pathway in standard-of-care-treated control GLASS samples, suggesting that this enrichment is specific to treatment with navtemadlin (**Fig. 2j**).

**Supplementary Figure 1.**
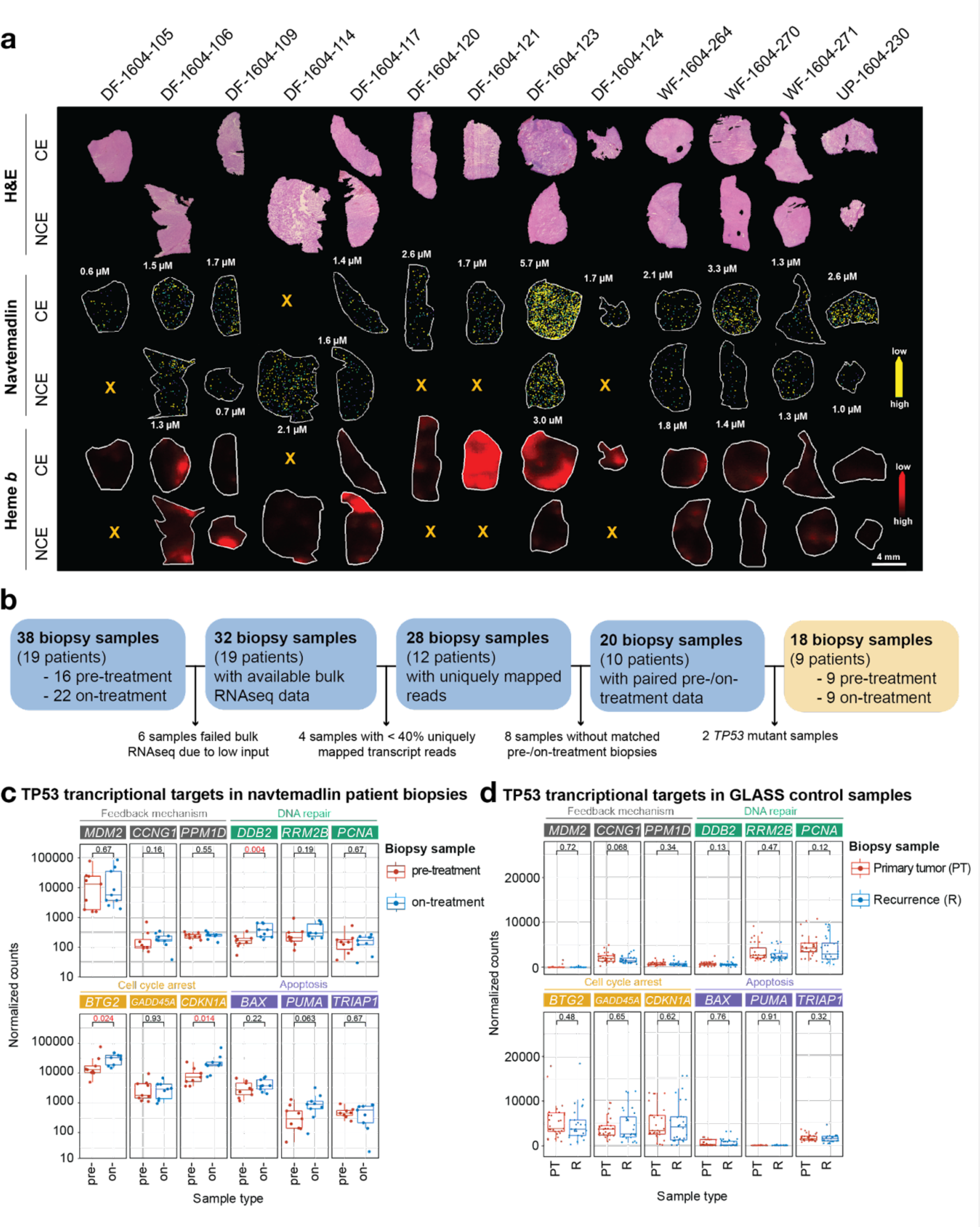
Evaluation of navtemadlin’s pharmacodynamic response in patient tumors. **(a)** Matrix Assisted Laser Desorption Ionization (MALDI) mass spectrometry imaging of navtemadlin and heme B from clinical trial specimens acquired from MRI contrast enhancing and non-enhancing tumor regions**. (b)** Selection of matched pre- and on-treatment *TP53* wild-type patient samples for bulk RNA sequencing. A total of 18 samples (yellow box) were chosen for downstream analyses. **(c)** Expression of TP53 transcriptional targets in navtemadlin-treated (blue) vs pre-treatment (red) patient biopsies. **(d)** Expression of TP53 transcriptional targets in primary tumor (red) and recurrence (blue) GLASS control samples.

### Cellular effects of navtemadlin treatment in patient-derived GBM models

Despite the upregulation of TP53 transcriptional targets, we did not observe evidence that p53 pathway inactivation contributed to tumor recurrence. Direct inactivation of p53 by mutation or deletion is a known resistance mechanism to MDM2 inhibition(16), so we sequenced *TP53* in three post-treatment tumors obtained from three patients who underwent biopsies post-recurrence. We detected no mutations or deletions and no upregulation of p53 by IHC (**Fig. 3a**).

**Figure 3.**
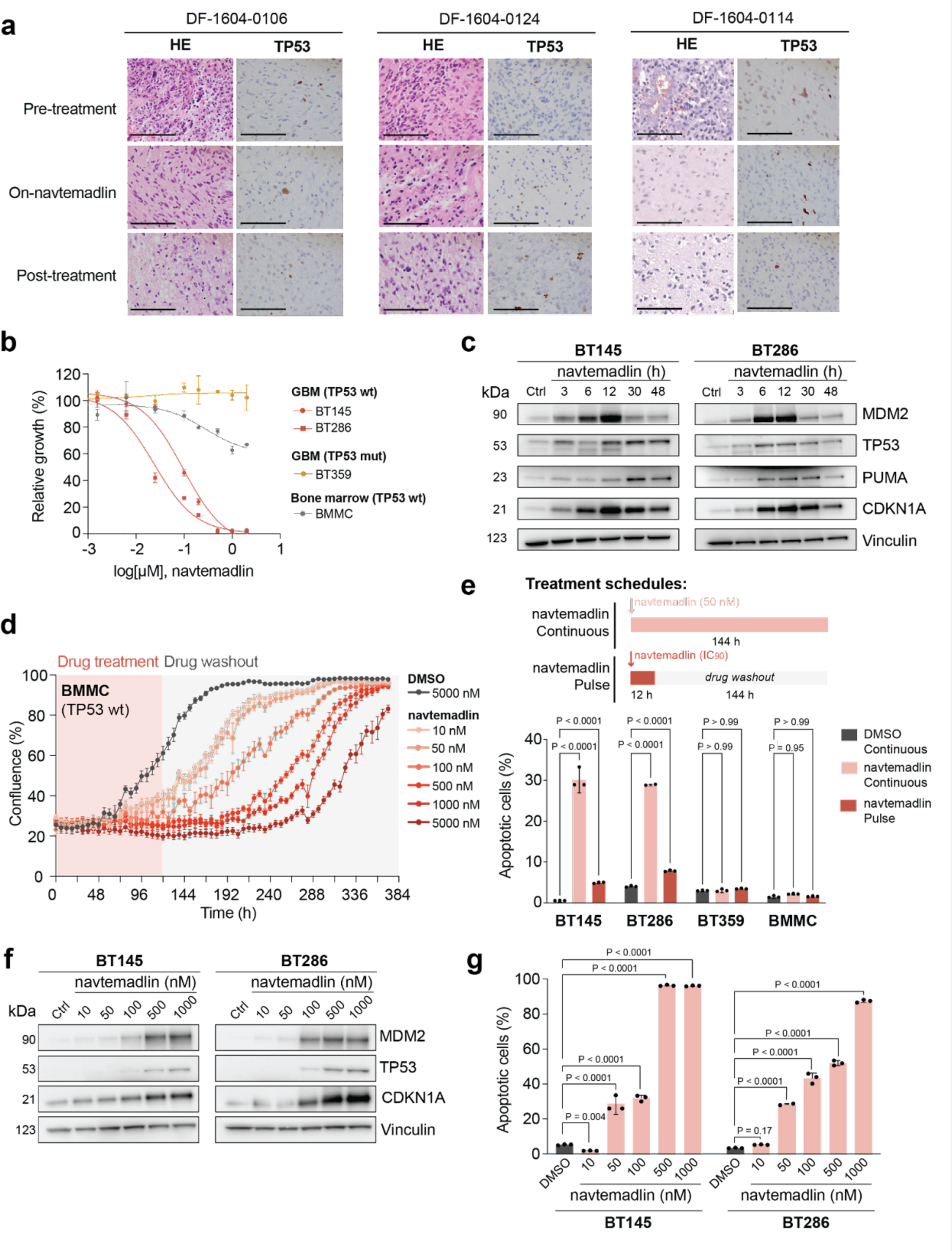
Effect of clinically relevant doses of navtemadlin on cell death. **(a)** Immunohistochemistry of p53 in three patients with available pre-treatment, on-treatment, and post-treatment biopsy samples. Scale bar = 100 μm. **(b)** Dose response curves showing sensitivity of *TP53* wild-type cells (BT145 and BT286; red), *TP53* mutant cells (BT359; yellow) and bone marrow cells (BMMC; gray) to navtemadlin. **(c)** Immunoblotting of BT145 and BT286 cells treated with 50 nM navtemadlin over time. Vinculin was included as a loading control. **(d)** Growth of BMMCs in response to navtemadlin treatment. Treatment with 50-5000 nM navtemadlin induces cell growth inhibition (red box), a phenotype that can be reversed when drug is washed out from the media (gray box). **(e)** Effect of continuous (light red) and single pulse (dark red) navtemadlin treatment modalities on cell death. For the single pulse modality, tested navtemadlin concentrations were: 100 nM in BT145, 75 nM in BT286, and 500 nM in BT359 and BMMCs. Apoptosis was quantified by annexin V/PI stainings. Data analyzed by Two-way ANOVA. **(f)** Immunoblotting of BT145 and BT286 cells treated with increasing concentrations of navtemadlin over 72h. Vinculin was included as a loading control. **(g)** Effect of increased navtemadlin treatment on tumor cell death. Apoptosis was quantified by annexin V/PI stainings after 72h of continuous drug treatment. Data analyzed by Two-way ANOVA.

To understand how tumors could recur in the absence of direct inactivation of *TP53*, we explored the effects of clinically relevant concentrations of navtemadlin on cell growth, viability and p53 pathway activity in both *TP53* wild-type (BT145 and BT286) and *TP53*-mutant (BT359) patient-derived neurosphere lines. First, we generated dose-response curves at 72 h of treatment, which showed the *TP53* wild-type cells to be significantly more sensitive than the *TP53*-mutant line (**Fig. 3b**). However, we did observe differences between BT145 and BT286. For example, at the concentration of 50 nM navtemadlin (representative of the lower drug concentration range found in the 120 mg cohort), we observed a >60% decrease in viability for BT286 cells but minimal response for BT145. Protein expression of PUMA (which is involved in apoptosis) but not CDKN1A, correlated with this response. With 50 nM navtemadlin, we detected PUMA activation by 6 hours into treatment in BT286 cells, but BT145 cells did not exhibit substantial increases in PUMA until 12 hours after treatment (**Fig. 3b**). In contrast, CDKN1A upregulation occurred in both *TP53* wild-type cell lines to a near-equal extent within 6 h of treatment.

Our results supported the concept of a therapeutic window between *TP53* wild-type gliomas and normal cells. Given the risk of MDM2 inhibition to bone marrow and other tissues with rapidly dividing cells (14,17,18), we separately included an immortalized bone marrow mesenchymal cell line (BMMC; T0523) in our 72 h navtemadlin dose-response study (**Fig. 3c**). BMMCs exhibited intermediate sensitivity to MDM2 inhibition, with large decreases in viability observed only with >1000 nM navtemadlin. Moreover, this decrease in viability resolved with removal of the drug, even at very high doses of navtemadlin. We treated these BMMCs for 5 days with increasing concentrations of navtemadlin and monitored cell confluency over time by live-cell imaging. Treatment with as little as 10 nM navtemadlin caused growth inhibition, with the strongest phenotypes being observed at >500 nM (**Fig. 3d**). However, when we washed-out drug from the media, we observed a resumption of growth to confluency among all conditions up to 5000 nM—a 35-fold higher concentration of navtemadlin compared to the 140 nM median achieved in the resected brain tumors from patients treated with the higher 240 mg dose. At six days into treatment with continuous doses of 50 nM navtemadlin, flow cytometry for annexin V and PI detected increased apoptosis relative to DMSO controls in both *TP53* wild-type lines, but not the *TP53*-mutant BT359 line or the BMMC line (**Fig. 3e; Supp. Fig. 2a-b**). These data suggest that the observed suppression in growth among BMMC cells reflected growth inhibition rather than apoptosis, further supporting evidence of a therapeutic window for *TP53* wild-type gliomas.

However, we did not observe complete ablation of *TP53* wild-type GBM cells at concentrations lower than 500 nM. To further understand the dose of navtemadlin that would be required for increased p53 pathway activation and cell death, we performed a comprehensive functional characterization of our *TP53*-wildtype preclinical models following treatment with a wide range of inhibitor concentrations. First, we measured levels of CDKN1A protein levels by immunoblotting as a marker of p53 pathway activation (**Fig. 3f**), and determined that at least 500 nM navtemadlin is required for robust protein expression. At this concentration, we additionally observed a clear increase in MDM2 and p53 protein levels, in agreement with navtemadlin’s role as a MDM2-p53 antagonist (14,19). By performing flow cytometry-based apoptosis analysis of these cells, we confirmed that at least 500 nM navtemadlin is required for achieving at least 60% of cell death by apoptosis. In fact, complete cell death in both cell line models was only achieved when treating with 1000 nM navtemadlin (**Fig. 3g; Supp. Fig. 2c**).

We considered two alternatives to increase rates of cancer cell death following navtemadlin treatment. First, we asked whether a pulsed high dose would induce more cell death than continuous dosing. A prior study suggested that pulsed IC_80_ doses of an MDM2 inhibitor (siremadlin; HDM-201) enhanced apoptotic signaling over continuous dosing (20). We tested single pulses at the following navtemadlin concentrations: 100 nM for BT145, 75 nM for BT286, and 500 nM for BT359 and BMMC cells. Surprisingly, pulsed dosing at IC_90_ levels did not increase the fraction of apoptotic cells in any of our models at six days following a 12 h treatment pulse (**Fig. 3e**). We conclude that continuous low-dose navtemadlin treatment causes more GBM cell death than a pulsed high-dose regimen.

### Dual treatment with navtemadlin and TMZ increases tumor cell death rates in GBM models

As a second alternative to continuous dosing of navtemadlin monotherapy, we explored whether we could enhance its apoptotic effects by applying it in combination with a DNA-damaging agent. Because the ongoing, second phase of this trial is already testing navtemadlin in combination with radiation, we decided to explore combined treatment with TMZ. TMZ is an alkylating agent that preferentially damages DNA in GBMs lacking expression of the DNA repair protein MGMT, usually due to *MGMT* promoter methylation, and is a component of first-line treatment for these tumors. We hypothesized that the replication stress caused by TMZ-induced DNA damage would sensitize cells to p53 pathway activation by navtemadlin. Indeed, TMZ and navtemadlin have been found to have non-overlapping mechanisms of action and have been described as synergistic in the context of traditional (non-neurosphere) glioma cell lines (21,22).

We found that navtemadlin and TMZ synergize in neurosphere cultures of *MGMT*-methylated GBMs, leading to high rates of apoptosis with an evident therapeutic window relative to bone marrow-derived controls. Treatment with both agents for 160 h caused a reduction in cell growth (*P* < 0.0001) for BT145 and BT286 (*TP53* wild-type and *MGMT*-methylated) but not BT359 (*TP53* mutant and *MGMT* unmethylated) cells (**Figure 4a; Supp. Fig. 2d**). In sensitive cell lines, a highest single agent (HSA) model found dual treatment with navtemadlin and TMZ to be synergistic for growth inhibition (**Supp. Fig. 2e**). The combined treatment led to increased protein levels of MDM2 and p53 at late timepoints (>30 h), relative to navtemadlin alone. CDKN1A levels were modestly increased in BT286 with combination treatment, but not in BT145 (**Figure 4b**). Flow cytometry for Annexin V and PI confirmed increased rates of apoptosis in BT145 and BT286 *TP53* wild-type cells following dual treatment. In BT286 cells, where sensitivity to TMZ treatment was the highest, addition of navtemadlin increased cell death across the tested concentration range (**Figure 4c**). In contrast, less than 5% cell death was observed in BT359 *TP53* mutant tumor cells or BMMCs treated with this combination, suggesting it retains a therapeutic window for the treatment of *TP53* wild-type and *MGMT* methylated GBMs (**Figure 4c; Supp. Fig. 2e**).

**Figure 4.**
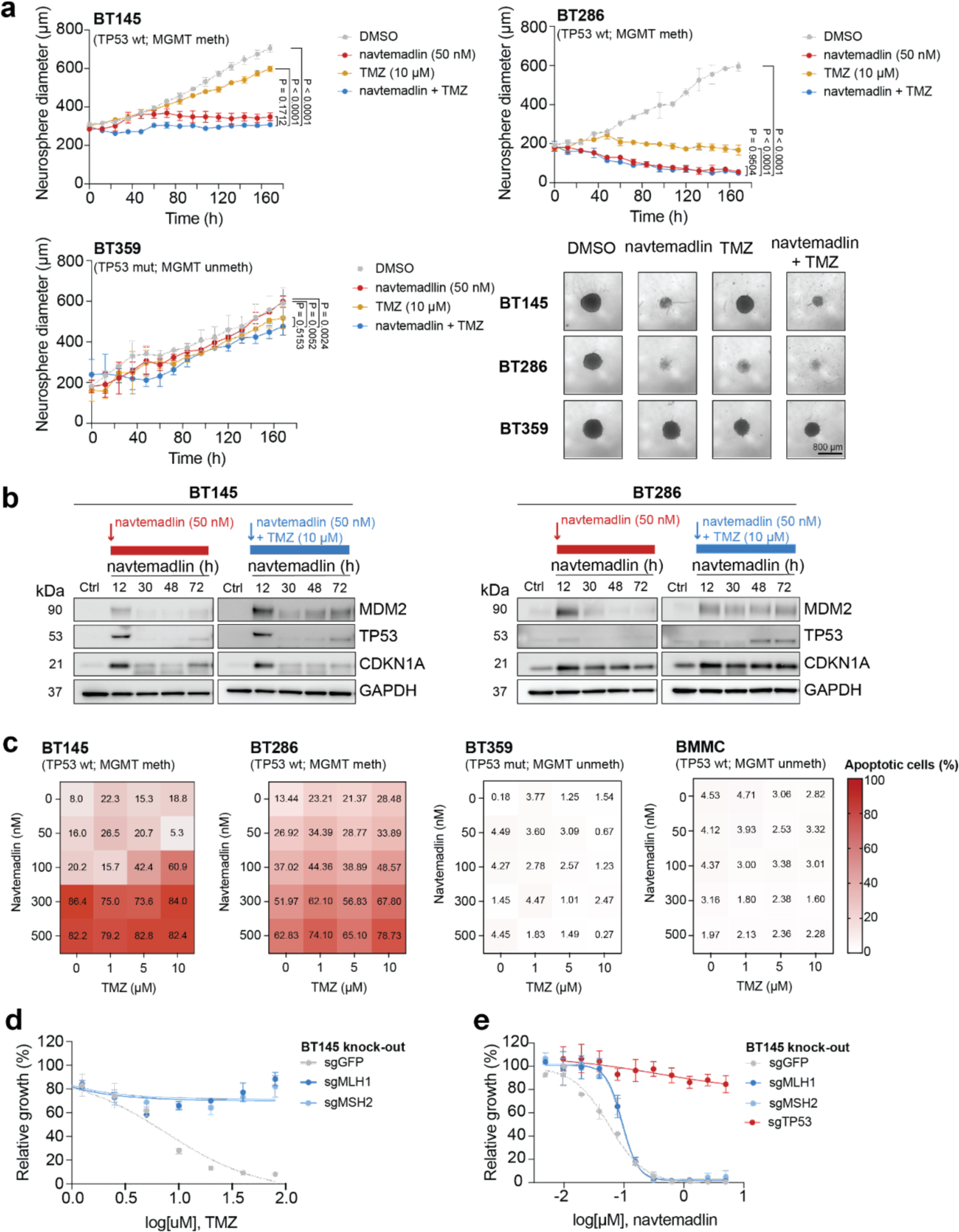
Effect of combination therapy with navtemadlin and TMZ on cell death rates. **(a)** Growth of *TP53* wild-type BT145 and BT286 cells when treated continuously with navtemadlin and TMZ as monotherapy or in combination. Growth of *TP53* mutant BT359 cells was performed as control. Mean and standard deviation of three technical replicates shown. Data analyzed by Two-way ANOVA. Neurosphere diameter was quantified by live imaging over the course of 160 h for cells treated with navtemadlin (red), TMZ (yellow) or combination therapy (blue). Representative images of TP53 wild-type (BT145 and BT286) and *TP53* mutant cells treated for each condition are shown to the right. Scale bar: 800 μm. Data analyzed by Two-way ANOVA. **(b)** Immunoblot of BT145 (left) and BT286 (right) cells treated with navtemadlin as monotherapy or in combination with TMZ for up to 72h. **(c)** Effect of navtemadlin and TMZ combinations on cell death. **(d-e)** Dose response curve showing sensitivity of BT145 *MLH1* and *MSH2* knock-out cells to treatment with **(d)** TMZ and **(e)** navtemadlin. A GFP sgRNA was used as CRISPR/Cas9 targeting control. Mean and standard deviation of three technical replicates shown.

Beyond attaining synergy, a major goal of combination therapy is to overcome resistance by adding treatments with non-overlapping mechanisms of action. Among MGMT-methylated GBMs, a primary resistance mechanism to TMZ treatment is loss of genes involved in DNA mismatch repair (MMR) (23). We engineered isogenic versions of the *MGMT* methylated BT145 GBM neurospheres both with and without each of the mismatch repair components MLH1 and MSH2. As expected, MMR loss led to TMZ resistance (**Fig. 4d**). However, genetic ablation of MMR genes led to only a slight decrease in sensitivity to navtemadlin, and these cells remained far more sensitive than an isogenic *TP53*-null BT145 cell model (**Fig. 4e**). We therefore conclude that resistance to TMZ does not necessarily lead to resistance to navtemadlin.

**Supplementary Figure 2.**
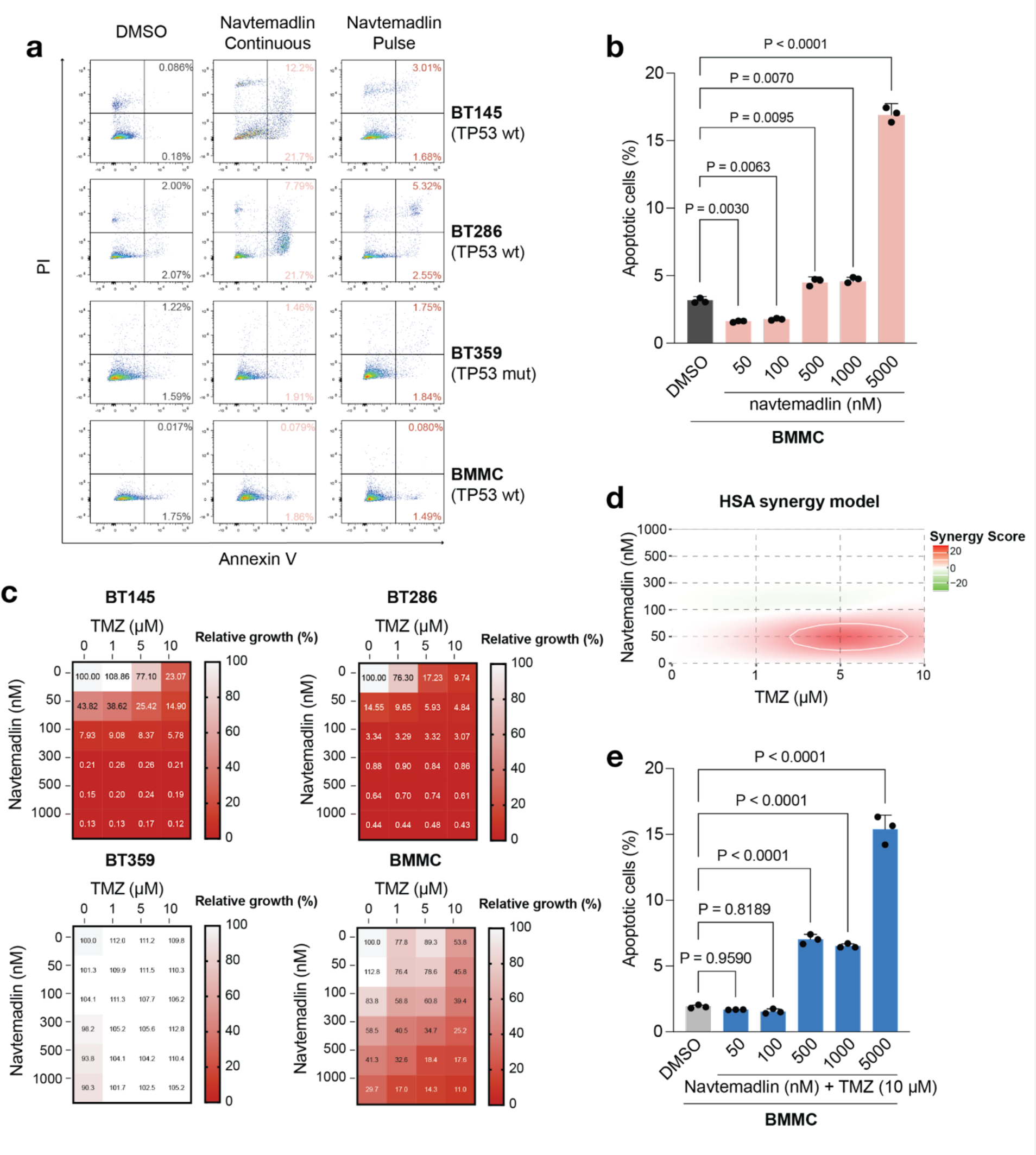
Effect of navtemadlin as monotherapy or in combination with TMZ. **(a)** Effect of continuous and single pulse navtemadlin treatment modalities on cell death. Representative flow cytometry stainings for the quantification shown in Fig. 4d. Data analyzed by Two-way ANOVA. **(b)** Percentage of cell death by apoptosis in bone marrow mesenchymal cells (BMMC) treated with navtemadlin. Quantification was done by flow cytometry-based annexin V and PI stainings. **(c)** Growth of *TP53* wild-type (BT145 and BT286) and mutant (BT359) GBM cells, as well as BMMC, in response to combination therapy with navtemadlin and TMZ for 5 days. Cell growth was acquired by CellTiterGlo luminescent measurements and quantified relative to DMSO control. **(d)** Synergy scores for navtemadlin and TMZ using a highest single agent (HSA) reference model. The analysis was performed in SynergyFinder [https://synergyfinder.fimm.fi/]. **(e)** Percentage of cell death by apoptosis in bone marrow mesenchymal cells (BMMC) treated with navtemadlin and TMZ. Quantification was done by flow cytometry-based annexin V and PI stainings.

### Treatment with navtemadlin induces adaptive changes in cell state

Apart from genetic resistance, we investigated whether adaptive changes in cell state contribute to navtemadlin resistance. A prior study has shown that inducing differentiation through exogenous expression of the oligodendrocyte transcription factor *OLIG2* decreases sensitivity to MDM2 inhibitors (24). We first interrogated our bulk RNA sequencing data from the nine patients with *TP53* wild-type tumors that were treated with navtemadlin. Relative to pre-treatment samples, on-treatment samples exhibited significant upregulation (adjusted p-value < 0.05) of genes associated with the GO term “oligodendrocyte differentiation” (GO:0048709), including *PLP1*, *NKX6-2*, *OPALIN*, *TPPP*, *HDAC11*, *DAAM2*, *MAL*, *CNP*, *SLC45A3*, *KCNJ10*, *ABCA2*, and *CNTNAP* (**Fig. 5a**). We also used signatures associated with developmental and oncogenic programs developed in a prior analysis of high-grade gliomas (25) to interrogate these RNA sequencing data using GSEA. On-treatment biopsies were enriched for astrocyte-like (AC-like) and oligodendrocyte-like (OC-like) differentiation programs (p < 0.001) but depleted of the “cell cycle” signature (p < 0.001) (**Fig. 5b**). In contrast, we did not observe enrichment for these signatures when analyzing GLASS control samples (p=0.407 and p=0.690 for AC-like and OC-like, respectively), nor could we detect a decrease in cycling cells (p=0.551) (**Fig. 5c**).

**Figure 5.**
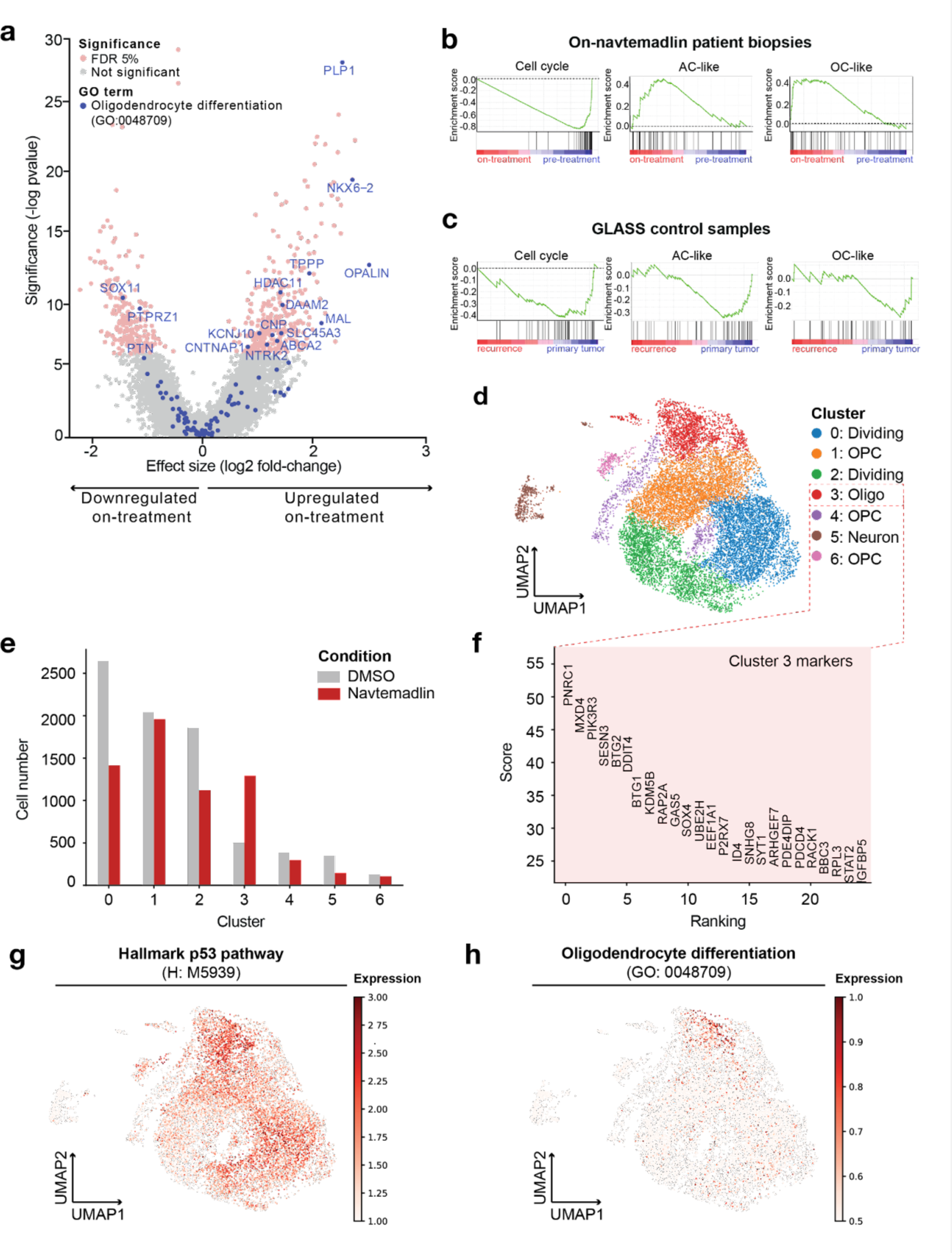
Navtemadlin induces differentiation of glioma cells. **(a)** Genes that become significantly deregulated with navtemadlin treatment (red; FDR cutoff 5%) include regulators of oligodendrocyte differentiation (blue). Gene Set enrichment analysis in **(b)** patient biopsies and **(c)** GLASS control samples using the cell state signatures described in (25). **(d)** UMAP unsupervised clustering of BT145 cells treated with 50 nM navtemadlin or DMSO control. **(e)** Number of cells per condition present in each cluster. **(f)** Markers defining cluster 3 when compared to remaining clusters. **(g-h)** Expression of **(g)** Hallmark p53 pathway and **(h)** oligodendrocyte differentiation gene signatures in BT145 cells and projection on the UMAP.

To get better resolution into the effects of navtemadlin on the proportion of different cell types and pathways active in each, we also performed single-cell RNA sequencing of BT145 cells treated with 50 nM navtemadlin vs. DMSO control for 15 hs (**Fig. 5d**). Using the Leiden algorithm (26) we detected seven clusters (Clusters 0-6), four of which represent oligodendrocyte precursor (Clusters 1, 4, and 6) and more differentiated oligodendrocyte cells (Cluster 3), one expressing neuronal markers (Cluster 5), and two clusters expressing high levels of histone and cell division genes (Clusters 0 and 2). MDM2 inhibition led to a decrease in dividing cells (Clusters 0 and 2) as well as an increase in oligodendrocyte-like cells (Cluster 3) (**Fig. 5e**). Cluster 3 was indeed characterized by high expression of genes involved in p53 signaling (e.g. *MXD4*, *DDIT4*, *BTG1*, *BTG2*, *RACK1*), apoptotic cell death (e.g. *BBC3*, *P2RX7*, *PDCD4*, *PIK3R3*, *SOX4*, *ARHGEF7*), and development (e.g. *KDM5B*, *ID4*, *SYT1*) (**Fig. 5f**). When we projected signatures of p53 pathway activation on UMAP visualizations of our data (**Figs. 5g-h**), we identified enrichment in cluster 3 (p=2.2×10^−16^; **Fig. 5g**). Interestingly, this same subset of cells was marked by high expression of the oligodendrocyte differentiation signature used to analyze our on-treatment human tumor biopsies (p=2.2×10^−16^) (**Fig. 5h**). We therefore conclude that navtemadlin induces a pharmacodynamic response that is detectable at the single-cell level and involves increased levels of an oligodendrocyte differentiation signature.

## Discussion

In this surgical window of opportunity trial in recurrent GBM, the primary endpoint of drug penetration into tumor tissue was met in both the 120 mg cohort and 240 mg cohort. Navtemadlin was tolerable with expected toxicities (6). The study also showed that navtemadlin penetrates both CE and NCE tumor tissue and exerts a pharmacodynamic effect in tumor tissue. Although the study was not powered to evaluate survival outcomes, PFS and OS were similar to historical controls (PFS of 1.5-6 months and OS of 2-9 months) (27).

Increased use of tissue sampling during glioma clinical trials has recently been advocated (28); this study highlights its utility. In general, lack of response may reflect inability of the treatment to reach cancer cells, lack of a direct pharmacodynamic response on the therapeutic target, lack of cancer cell killing, acquisition of resistance among a minor cell population, or a combination of these factors. We were able to use on- and post-treatment biopsies, combined with an *in vitro* evaluation of navtemadlin in representative cell lines, to address all of these possibilities. By detecting a pharmacodynamic response without sufficient cell killing, we can focus on the most promising paths to improving the use of MDM2 inhibitors like navtemadlin in future trials.

Although apoptosis markers BAX and PUMA were expressed in post-treatment biopsies, increased activation of apoptosis is likely necessary for durable clinical benefits. One approach might be to increase on-target inhibition: *in vitro* studies with representative *TP53* wild-type GBM neurosphere models showed dose-dependent increases in apoptosis and the upregulation of p53-mediated pathways in multiple populations defined by single-cell RNAseq, with nearly 100% cell death at 1000 nM concentrations.

We also observed adaptive changes in the cell differentiation state with 50 nM navtemadlin, as evidenced by the upregulation of genes involved in oligodendrocyte differentiation, along with a decrease in dividing cell clusters. Tumor RNAseq also demonstrated that genes associated with oligodendrocyte differentiation were among the significantly affected genes in the resected tumor tissue after ∼30 hours of navtemadlin treatment, with enrichment scores for both AC-like and OC-like gene sets. High-grade gliomas can indeed adapt to therapeutic pressures by acquiring genetic or transcriptional alterations that modify their cell state (25,29–31). Decreased expression of GBM stemness markers has been observed in patient-derived *in vitro* cell line spheroids treated with navtemadlin (10). However, this represents the first clinical study in any disease to detect such adaptive changes in human tumor tissue treated with MDM2 inhibitors. Prior studies have indicated that progenitor-like cell states among GBM cells promote resistance to TMZ, radiation, and other treatment approaches such as immunotherapy (32–35). This contrast–the relative resistance of stem-like cells to standard radiation and TMZ, versus the induction of signatures of more differentiated cells by p53 reactivation–supports the concept of combining these treatment modalities.

Such combinations of DNA-damaging treatments with targeted p53 pathway activation may also increase rates of cancer cell death. Our preclinical analyses in GBM models revealed that the lower range of clinically relevant navtemadlin concentrations in patient CE and NCE tissues activates the p53 pathway–including apoptotic signaling–but is insufficient to fully kill the cancer cell population *in vitro*. A strategy to overcome partial cell death is increasing the dose of the MDM2 inhibitor while accounting for toxicity in normal treatment-sensitive tissues. Alternative dosing schedules, such as single high-doses pulses, have been reported to increase apoptosis in *TP53*-wildtype, *MDM2*-amplified osteosarcoma cells with a different MDM2 inhibitor (20). We did not observe increases in cell death rates at six days when pulsing navtemadlin for 12 h in GBM cell lines, but our results suggest that higher concentrations of drug (>500 nM) may be required to achieve complete tumor cell killing. A second strategy is to test more potent MDM2 inhibitors with increased brain penetrance, such as the MDM2 inhibitor BI-907828 (11), which acts at a picomolar concentration i*n vitro* and is currently being evaluated in patients with advanced or metastatic *TP53*-wildtype solid tumors (NCT03449381). Thirdly, it will be critical to consider combination therapy with agents that exhibit non-overlapping mechanisms of action with MDM2 inhibition. Here, we have shown the potential of combining navtemadlin with TMZ in *MGMT* methylated tumors. One major concern is potential induction of thrombocytopenia by both navtemadlin and TMZ; however, bone marrow cells were much more tolerant of the combination than glioma cells *in vitro*. In this regard, the bone marrow cell cultures recovered from cell cycle arrest and resumed proliferation following discontinuation of *in vitro* treatment, rather than showing commitment to apoptotic cell death. Moreover, myeloid suppression is a standard feature of widely used treatments for other cancers (36), and can be managed with a combination of cytokine-based myeloid stimulation and transfusions. The combination approach thus has the potential to extend the population of patients eligible for therapy with MDM2 inhibitors.

This study has provided evidence that navtemadlin’s effectiveness is enhanced with agents that increase replication stress. However, patients with unmethylated *MGMT* are unlikely to benefit from combinations that include navtemadlin with TMZ. Since RT also induces replication stress, we are now proceeding with the phase 1 component (NCT03107780), a dose escalation study combining navtemadlin with radiation in patients with newly diagnosed, *MGMT* unmethylated, TP53 wild-type GBM within Alliance.

## Data Availability

All data produced in the present study are available upon reasonable request to the authors

## Acknowledgments

This study was sponsored by the Adult Brain Tumor Consortium (ABTC) NIH/NCI UM1 CA137443, NIH grants U54-CA210180 (NYRA), P41-EB028741 (NYRA), U19CA264504, R01CA262462 (RB and KL), R0188228 (RB and KL), and Capital Award from the Massachusetts Life Sciences Center. The project described was also supported by the Analytical Pharmacology Core of the Sidney Kimmel Comprehensive Cancer Center at Johns Hopkins [NIH grants P30CA006973 and UL1TR003098, and the Shared Instrument Grant S10RR026824], the Gray Matters Brain Cancer Research Foundation, The Brain Tumour Charity, and the Swedish Research Council. The grant number UL1TR003098 is from the National Center for Advancing Translational Sciences (NCATS), a component of the National Institutes of Health (NIH), and the NIH Roadmap for Medical Research. Its contents are solely the responsibility of the authors and do not necessarily represent the official view of the NCATS or NIH.

## Methods

### Trial design

The phase 0 surgical cohort of Adult Brain Tumor Consortium (ABTC) protocol 1604 (NCT03107780) enrolled adult participants with recurrent glioblastoma (GBM) who were planned for surgical resection. Additional eligibility criteria included Karnofsky Performance Status (KPS) ≥ 60% as well as normal organ and marrow function. Any number of prior therapies including prior bevacizumab use (following a washout of 6 weeks from bevacizumab) was allowed on study. Use of enzyme-inducing antiepileptics, anticoagulants (other than low molecular weight heparin and prophylactic low dose warfarin), and tumor-treating fields was not allowed on study. Prior to surgery, participants were sequentially allocated to receive navtemadlin at either 120 mg daily or 240 mg daily for 2 days prior to surgery. Surgery was performed 3-6 hours following the last administration of navtemadlin. Blood samples for measurement of macrophage inhibitory cytokine-1 (MIC-1) levels were collected at baseline/screening, on the day prior to surgery prior to the first dose, and 24 hours post first dose but before the second dose on the day of surgery. Upon adequate recovery from surgery, participants whose tumors were *TP53* wild-type were eligible to continue navtemadlin at the recommended phase 2 dose for monotherapy (240 mg daily for 7 days every 3 weeks) until progression or unacceptable toxicity.

### Pharmacokinetics

Navtemadlin levels in brain tissue were measured by a validated liquid chromatography coupled with tandem mass spectrometry (LC-MS/MS) method over the range of 1-1,000 ng/mL (37). Descriptive statistics were used to summarize the pharmacokinetic results of contract and non-contrast enhancing brain tissue.

### Evaluation of brain drug distribution by Matrix Assisted Laser Desorption Ionization (MALDI) mass spectrometry imaging

Frozen tissue specimens were cryo-sectioned to 10 µm thickness and thaw-mounted onto indium tin oxide (ITO) coated glass slides. Consecutive sections were mounted onto microscopy slides, stained with hematoxylin and eosin (H&E), and imaged with a 10X objective (Zeiss Observer Z.1, Oberkochen, Germany). A matrix solution of 2,5-dihydroxybenzoic acid (160 mg/mL) in 70% methanol:30% trifluoroacetic acid (TFA), 1% dimethyl sulfoxide (DMSO) was applied onto tissue sections using a robotic sprayer (HTX Technologies, Chapel Hill, NC) using a two-pass cycle with flow rate of 0.18 mL/min, spray nozzle velocity of 1200 mm/min, nitrogen gas pressure of 10 psi, spray nozzle temperature of 75 °C, and a track spacing of 2 mm. Recrystallization was performed using 5% acetic acid solution at 85 °C for six minutes. Mass spectrometry imaging data was acquired using a timsTOF fleX mass spectrometer (Bruker Daltonics, Billerica, MA) in positive ion mode with a multiple reaction monitoring (MRM) method scanning across m/z 100-650. The mass spectrometry method was developed and optimized by direct infusion of a navtemadlin solution in the electrospray ionization (ESI) source for a transition of the precursor ion from 568.168 → 301.051 (collision energy 45 eV, 3 m/z isolation window). Data was analyzed and visualized using SCiLS Lab software (version 2021a premium, Bruker Daltonics, Billerica, MA) with a total ion current (TIC) normalization.

### Pharmacodynamics

For pharmacodynamic analysis from peripheral blood, a commercially available Human GDF-15 Quantikine ELISA kit (RnD Systems, Minneapolis, MN) was utilized to measure serum MIC-1 levels over the range of 23.4 to 1500 pg/mL with dilutions up to 1:50 (v/v) being validated. Since serum MIC-1 changes in a concentration-time like profile, samples lacking a paired pre-dose baseline were not utilized in the analysis (38). This excluded six patients from the 120 mg and four from the 240 mg cohorts. The FCB analysis was performed on the pre-dose samples that were obtained on the day of surgery. Two baseline measurements were obtained to assess the variability in the baseline to ensure robustness in the assessment as to whether navtemadlin altered MIC-1 sufficiently. Descriptive statistics were used to summarize the results.

### Immunohistochemistry

An expert neuropathologist reviewed histological patient tumor samples in order to assess the WHO 2016 integrated diagnosis and to select the tumor areas for immunohistochemistry. Drug effects on tumor tissue were evaluated on formalin-fixed paraffin sections using Envision DAB staining system (DAKO). To assess drug modulation of the p53 pathway, the following antibodies were used: mouse monoclonal anti-p53 (Agilent, clone M7001, 1:100), mouse monoclonal anti-MDM2 (Millipore Sigma, clone OP46, 1:50), rabbit monoclonal anti-p21 (Cell Signaling, clone 12D1, 1:50), rabbit polyclonal anti-phospho-CDK1 (Thermo Fisher, clone PA5-85508, 1:500), rabbit monoclonal anti-Cleaved-Caspase 3 (Cell Signaling, clone 5A1E, 1:2000), and mouse monoclonal anti-Ki67 (Agilent, clone M7240, 1:100). Pathway response was assessed by quantification of the percentage of nuclei staining positively for each target using HALO AI digital pathology software (v3.6 Indica Labs, New Mexico, United States) on whole slide images. Artifacts areas such as air bubbles or ink marks were manually excluded. Nuclei Seg network detection algorithm was utilized to segment the boundaries of cell nuclei based on staining intensity and morphology. HALO Multiplex IHC v3.4 analyzer was used to identify positively stained nuclei according to specific staining intensity thresholds.

### Nucleic acid extraction and bulk RNA sequencing (RNAseq)

For the analysis of human biopsies, formalin-fixed paraffin-embedded (FFPE) tumor blocks were sectioned into 40 μM scrolls using a microtome. DNA and RNA were extracted simultaneously from each sample using the AllPrep DNA/RNA FFPE Kit (80234, Qiagen). Prior to nucleic acid extraction, paraffin was dissolved using a deparaffinization solution (19093, Qiagen). To extract RNA from BT145 and BT286 cell lines, two million cells were collected after treatment with 50 nM navtemadlin for 30 h. Next, RNA was isolated using the RNeasy Mini Kit with an on-column DNase digestion (79254, Qiagen). Resulting RNA concentrations were quantified with a Nanodrop 2000 Spectrophotometer (ND-2000, Thermo Scientific), and up to 500 ng were submitted for total RNA sequencing on an Illumina NovaSeq instrument at The Molecular Biology Core Facilities of Dana-Farber Cancer Institute (USA). Here, samples were subjected to rRNA depletion using the QIAseq FastSelect -rRNA HMR kit (334385, Qiagen) and sequencing was performed to generate 50M 150 bp read pairs (100M reads) per sample. Quality control, alignment and initial analyses were performed with VIPER (39). For both human tumor samples and cell lines, differentially expressed genes were calculated with the DESeq2 pipeline (40). For the analysis of GBMs treated with standard-of-care, transcript counts and clinical data for the samples reported in (41) were downloaded from The Glioma Longitudinal Analysis (GLASS) consortium [https://glass-consortium.org]. Significantly dysregulated genes were chosen based on FDR ≤ 5% and Log_2_ Fold-change ± 1 cutoffs.

### Pathway analysis

Count matrices from sequenced tumors and cell lines were subject to Gene Set Enrichment Analysis using the GSEA 4.2.3 software implementation. To determine cellular effects associated with treatment, navtemadlin treated samples and GLASS controls were analyzed using the hallmark gene sets from the Human MSigDB Collection (42). Enrichment analysis for cell differentiation signatures was conducted using the cell cycle, astrocytic differentiation (AC-like), oligodendrocytic differentiation (OC-like) and OPC-like programs described in (25). To assess p53 pathway activation in BT145 and BT286 patient-derived neurosphere models, we first identified differentially expressed genes in cells treated with 50 nM navtemadlin for 30 h relative to DMSO control. We detected 390 and 132 dysregulated genes in BT145 and BT286 cells, respectively, with 125 genes (31 upregulated and 96 downregulated) being commonly dysregulated in both cell lines. We then used these 125 genes to construct custom gene sets of p53 pathway upregulation and downregulation. Finally, we performed enrichment analysis of navtemadlin treated human tumors and GLASS controls using these gene sets.

### Single-cell RNAseq and analysis

All single-cell data was analyzed in Python 3.9.7 using Scanpy 1.7.0, Matplotlib 3.6.2, and Pandas 1.4.3. Total RNA read counts were normalized to 10,000 to control for variability in sequencing depth between cells. The data was then log transformed and scaled to unit variance while clipping logread counts to 10 to reduce the effect of outliers. Unhealthy cells with mitochondrial content greater than 20% were filtered out as well as cells expressing fewer than 200 genes. Finally, we included only highly variable genes and genes belonging to our gene sets of interest in our analysis to reduce noise from housekeeping genes. Genes are deemed highly variable using the ‘*highly_variable_genes*’ function in Scanpy with default settings which uses a measure of the variance of a gene across cells relative to its average expression. All plots were generated using the Scanpy and Matplotlib libraries. To visualize the data, we reduced the data using principal component analysis, with the *arpack* singular-value decomposition solver, and used the top 40 principal components to produce a neighborhood graph with a hard cutoff at 10 neighbors using the ‘*pca*’ and ‘*neighbors*’ functions respectively. This allowed us to embed the data into 2 dimensions into a uniform manifold approximation and projection (UMAP) plot using Scanpy’s ‘*umap*’ function. Pathway activation was scored based on a cell’s average expression of constituent genes in a pathway. Clustering was performed by Scanpy’s ‘*leiden*’ function(26), at a resolution of 0.2, which is a hierarchical algorithm that recursively merges communities of cells into progressively larger nodes.

### Cell culture

The patient-derived neurosphere cell lines BT145, BT286 and BT359 were obtained from the Center for Patient Derived Models (CPDM) at Dana-Farber Cancer Institute. Human bone marrow mesenchymal cells (BMMCs) immortalized with hTERT were commercially acquired (T0523, Abm). Neurosphere models were grown in Tumor Stem Medium (TSM), consisting of Neurobasal-A (10888022, Gibco), DMEM/F-12 (11330057, Gibco), Sodium Pyruvate (11360-070, Life Technologies), HEPES (15630-080, Life Technologies), Glutamax (35050-061, Life Technologies), non-essential amino acids (11140-050, Gibco) and 1% penicillin-streptomycin (15140-122, Life Technologies), and supplemented with B-27 (17504044, Gibco) and a growth factor mix consisting of 0.2% heparin (7980, Stem Cell), 20 g/mL bFGF (78002.2, Stem Cell) and 20 g/mL EGF (78006.2, Stem Cell). BMMCs were grown adherently in plastic coated with Geltrex (A1413302, Fisher Scientific), and cultured in RPMI 1640 medium (11875093, Gibco) supplemented with 10% fetal bovine serum (FBS; Gemini Bio, Cat. #100-800), 1% penicillin-streptomycin and 10 M hydrocortisone (Sigma Aldrich, Cat. #H0888-5G). HEK293T cells were grown in DMEM (12491, Gibco) supplemented with 10% FBS. All cell lines were maintained at 37 °C in 5% CO_2_, examined for mycoplasma contamination using the MycoAlert Mycoplasma Detection Kit (LT07-518, Lonza) and subjected to STR profiling at the Molecular Diagnostics Laboratory of Dana-Farber Cancer Institute.

### Generation of mismatch-repair deficient and TP53-null cells

To generate virus, Lipofectamine 3000 (L3000001, Thermo Fisher) reagents were used to transfect HEK293T cells with 10 μg psPAX2 (12260, Addgene) and 1 μg VSVG (12259, Addgene) viral packaging plasmids, as well as 10 μg of the pLX-311-Cas9 lentiviral vector (118018, Addgene) or the all-in-one pXPR_BRD051 vector containing guide RNAs targeting *TP53* (GAAGGGACAGAAGATGACAG), *MSH2* (ATTCTGTTCTTATCCATGAG) or *MLH1* (GAGATGATTGAGAACTGGTA) genes (obtained from the Genetic Perturbation Platform at the Broad Institute of MIT and Harvard). Virus was harvested 48 h later with the use of a 0.45 micron syringe filter. For transduction of the different lentiviral plasmids, BT145 cells were seeded at a density of 2 million cells per well in a 12-well plate (353225, Corning). Virus was added together with 5 μg mL^-1^ of polybrene, and cells were centrifuged at 2,000 rpm for 2 h at 30 °C. BT145 cells were first transduced with the Cas9 plasmid, and selected with 10 μg/mL blasticidin (A1113902, Gibco) for 7 days. To generate knock-out cells, BT145 Cas9-expressing cells were transduced again with pXPR-052 virus containing guide RNAs against the desired gene, and selected with 100 μg/mL hygromycin (10687010, Gibco) for 10 days.

### Immunoblotting

Pellets from three million cells were lysed with RIPA lysis buffer (9806S, Cell Signaling Technology) along with a protease and phosphatase inhibitor cocktail (PI-285, Boston BioProducts) and PMSF (8553S, Cell Signaling Technology). Lysates were incubated for 45 min on ice with pulse-vortexing in 15 min intervals and centrifuged at 13,000 rpm for 10 min in 4 C. Protein concentrations were then quantified by BCA against a bovine serum albumin standard. Absorbance was measured by incubating protein samples and BCA standards with Pierce A/B colorimetric reagents (23227, ThermoFisher). A total of 30 μg protein was loaded for each sample in a NuPAGE 4-12% Bis-Tris gel (NP0335BOX, Invitrogen) and run with 1X NuPAGE MOPS Running buffer (NP001, Life Technologies) for 1.5 h at 120V. For protein size comparison, the Cytiva full-range Rainbow molecular weight marker (RPN800E, Fisher Scientific) was used as standard. Following protein separation, a dry transfer was completed at 30V for 6 min with PVDF stacks in an iBlot 2 transfer instrument (IB24002, Invitrogen). After gel transfer, the membrane was blocked with 5% dry milk (1706404, BioRad) in TBS-T (T1680, Fisher Scientific) for 30 min. Primary incubation with antibodies against rabbit p53 (2527S, Cell Signaling Technology), rabbit PUMA (12450S, Cell Signaling Technology), rabbit CDKN1A/p21 (2947S, Cell Signaling Technology), rabbit MDM2 (86934S, Cell Signaling Technology) and rabbit GAPDH (5174S, Cell Signaling Technology) at a 1:1000 dilution occurred overnight at 4 °C. After washes with TBS-T, the membrane was incubated with an anti-rabbit secondary antibody (7074S, Cell Signaling Technology) at a 1:3000 dilution for 1 h at room temperature. For the detection of apoptosis, a primary apoptosis antibody cocktail (Ab136812, Abcam) was used at a 1:250 dilution followed by a HRP-cocktail at a 1:100 dilution. Signal was detected by chemiluminescence with SuperSignal West Pico (34095, Life Technologies) and Femto kits (34580, Life Technologies) in an ImageQuant LAS 4000 imager (GE Healthcare Life Sciences). When needed, membranes were stripped with Restore Plus Western Blot Stripping Buffer (46430, Thermo Fisher Scientific) for 15 min, washed with TBS-T, and blocked with 5% milk and TBS-T for 30 min. Blots were quantified using ImageJ 1.52k software (NIH, USA), where band intensity was normalized against the loading control and expressed relative to the untreated condition.

### Growth curves

Cell growth was tracked in real-time using the IncuCyte S3 Live Cell Analysis System (Sartorius). Mesenchymal bone marrow cells, which grow adherently, were seeded at a density of 5,000 per well in a clear 96-well plate (353075, Corning) precoated with 1X Geltrex (A1569601, Thermo Fisher). The next day, drug treatment was initiated and images were captured using a 10X objective every 6 h. Cell confluency was determined for each condition in six technical replicates. For neurosphere cell lines (BT145, BT286, and BT359), cells were seeded in ultra-low attachment round bottom 96-well plates (7007, Corning) at a density of 500 cells per well. After a centrifugation for 15 min at 200 *g*, the plates were inserted in the IncuCyte System and images were acquired every 6 h using the 4X objective. Neurosphere diameter was measured in ImageJ 1.52k (NIH, USA). Statistical analysis was performed in GraphPad Prism.

### Dose response curves and drug synergy analysis

Patient-derived neurosphere models (BT145, BT286, and BT359) and BMMCs were treated with the MDM2 inhibitor navtemadlin (HY-12296, MedChem Express) and temozolomide (TMZ; S1237, Selleckchem) as single agents or in combination. To assess growth relative to a DMSO (12611, Cell Signaling Technology) control, 5,000 cells per well were seeded in triplicates in a 96-well white bottom plate (136102, Thermo Scientific). Drug treatments with navtemadlin as monotherapy lasted >72 h, whereas experiments involving TMZ were extended to >120 h to allow for bioactivation of this prodrug. The CellTiter-Glo assay (G9681, Promega) was used to measure levels of ATP as a surrogate for cell viability. Briefly, the reagent mixture was added in a 1:1 ratio to the 96-well plate, which was then incubated at room temperature for 15 min. Luminescence measurements were acquired in a SpectraMax M5 plate reader (Associated Technologies Group) with an integration time of 500 ms. The data were analyzed in GraphPad Prism. Potential synergy between navtemadlin and TMZ was calculated using the web 3.0 version of SynergyFinder, where multi-dose viability data was uploaded for each cell line. The degree of combination synergy was quantified by the HSA model.

### Detection of apoptosis by flow cytometry

Cell death fractions following treatment with navtemadlin and/or TMZ were detected with the Alexa Fluor 488 annexin V/Dead Cell Apoptosis Kit (V13241, Thermo Fisher). For bone marrow cells, which are GFP-positive, an Alexa Fluor 647 annexin V antibody (A23204, Thermo Fisher) was used instead. Briefly, 1 million cells were pelleted and resuspended in 1X annexin-binding buffer (50 mM HEPES, 700 mM NaCl, and 12.5 mM CaCl_2_) containing 1:20 annexin V antibody and 1 μg/mL propidium iodide (PI). After a 15 min incubation at room temperature, cells were analyzed in a Fortessa 3 flow analyzer (BD Biosciences). Here, single cells were first gated using forward scatter (FSC) and side scatter (SSC) parameters, and were later classified in four quadrants based on whether they were negative for both stainings (viable), annexin v-positive (early apoptotic), PI positive (necrotic), or positive for both stainings (late apoptotic). The fraction of apoptotic cells was then calculated by adding the percentage of cells in the early and late apoptotic quadrants. Samples were measured in triplicates and analyzed in FlowJo 10.8.1.

