## Supplementary figures and images for "Surgical window of opportunity trial reveals mechanisms of response and resistance to navtemadlin (KRT-232) in patients with recurrent glioblastoma"

### Supplementary Figures

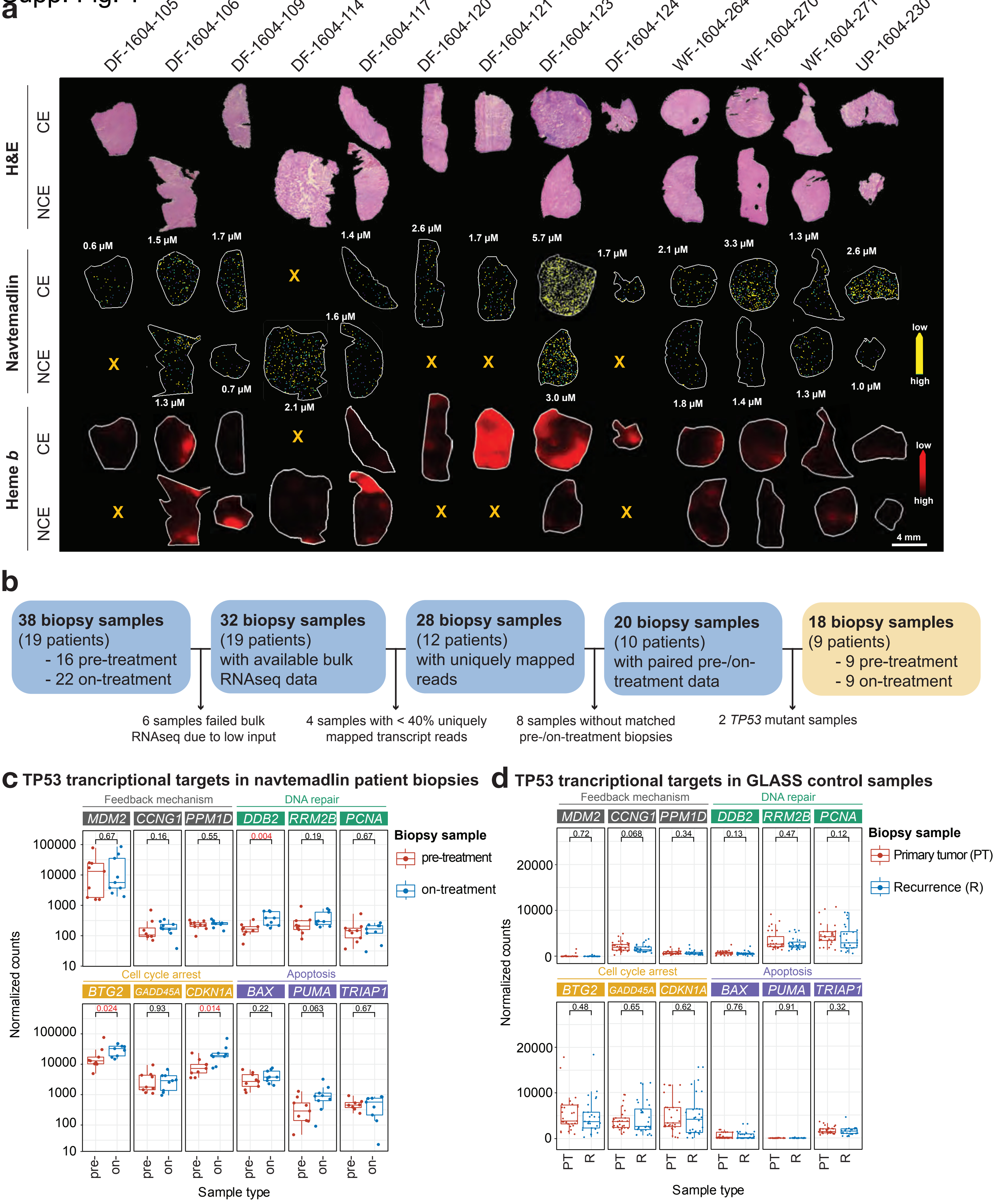

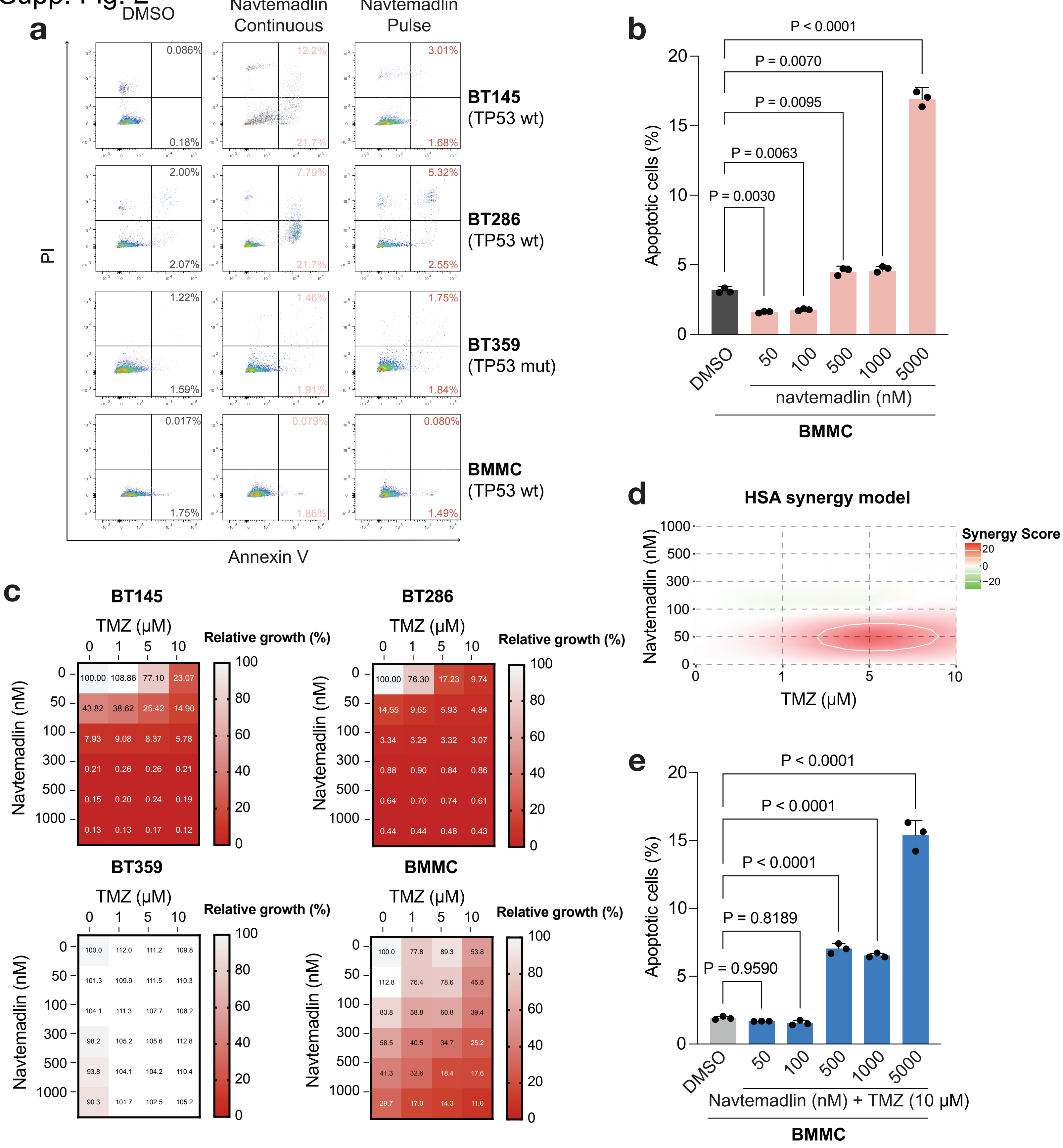
